# Partitioning lifespan inequality in hierarchical study designs with an application to global healthy longevity

**DOI:** 10.64898/2026.11.22.26368689

**Authors:** Hal Caswell

## Abstract

**Background:** Demographic inequality arises from two sources: heterogeneity among individuals subject to different rates, and stochastic variation among individuals subject to identical rates. Partitioning lifespan inequality into these components has been limited to heterogeneity in single factors or, very recently, factorial study designs with multiple factors. Methods for hierarchical designs have been lacking.

**Objectives:** The goal of this paper is to present a generally applicable method to partition inequality of lifespan (and other demographic outcomes) into components within and between levels in a hierarchical study design.

**Methods:** The analysis treats a heterogeneous population as a mixture distribution. Inequality is measured by the variance and partitioned into components between groups (e.g., geographical or economic regions), between sub-groups within groups (e.g., countries within regions), and stochasticity within sub-groups. The method is applied to global inequality in healthy longevity, using data for 194 countries from the Global Burden of Disease study, divided into four regions according to the World Bank income categories and into countries within each region. Results are compared for flat and population-weighted mixing distributions.

**Results:** The protocol for variance partitioning in hierarchical study designs provides variance among regions, among countries within regions, and within countries. It extends naturally to multiple levels (e.g., states within countries) and to factorial components (e.g., regions defined by combinations of income and education). When applied to global healthy longevity, the dominant source of inequality is stochastic variation within countries, which accounts for 85%-95% of the overall inequality. The population-weighted mixing distribution reduces the contribution of between-group variance.

**Contribution:** Some of the largest demographic data sets have a hierarchical organization. The methods presented here make it possible to partition inequality, measured by variance, in such studies.

## 1 Introduction: Chance or consequence?

The individuals in a cohort or a population differ among themselves in the outcomes of any demographic process. Of particular interest as outcomes are longevity or lifespan (or its various flavors, such as healthy longevity, which is our focus here), lifetime reproductive output, and lifetime availability of kin. This variation among individuals, often called ‘inequality,’ has important demographic and social consequences.

Where does this inequality come from? In Christopher Nolan’s 2026 film *The Odyssey*, the goddess Athena asks Odysseus, “When you look at death, do you see chance or consequence?” Her question^1^ highlights the most fundamental issue in asking where inequality comes from. Is it a consequence of some social, economic, or environmental factors (e.g., education, income, violence)? Or is it the result of chance, resulting from the repeated application of probabilities of survival and reproduction?

In a homogenous population, in which every individual is subject, at a given age, to the same mortality schedule, the inequality in longevity can only be due to chance. The standard life table analysis provides the probability distribution of lifespan (often denoted by *d*_*x*_) resulting from age-specific probabilities of death (often denoted by *q*_*x*_) that are applied uniformly to everyone. Differences among individuals arise solely due to chance (individual stochasticity; Caswell 2009).

In a heterogeneous population, however, individuals fall into different groups,^2^ each of which is subject to its own demographic rates. This creates a between-group component of inequality, as a consequence of those differences. Even so, each group is still characterized by a life table, which is applied identically to every individual within the group, producing a within-group component due to stochasticity.^3^

Heterogeneity has many potential sources. It may be due to observable external factors (e.g., income, education) or to unobservable latent factors (e.g., frailty), but in any case the factors produce differences among individuals in the risks they experience. In order to understand the effects of social and environmental factors on inequality, many authors have worked to partition the inequality into contributions of heterogeneity between groups and stochasticity within groups (e.g., Hoem and Jensen, 1982; Vaupel, 1988; Edwards, 2011; Hartemink and Caswell, 2018; Caswell, 2023).

### 1.1 Indices of inequality

There are many indices with which to measure inequality (e.g., Sen, 1997, Chap. 2). They have been extensively studied by economists, and some have been applied to demographic outcomes (e.g., Wilmoth and Horiuchi, 1999; Anand and Nanthikesan, 2000; Shkolnikov, Andreev, and Begun, 2003; van Raalte et al., 2012; Van Raalte and Caswell, 2013). In this paper I use the variance in the outcome (longevity in this case), because it can be partitioned into within-group and between-group components, thus revealing the balance between heterogeneity and stochasticity (see Section 2). The other decomposable index is the entropy-based Thiel index. The Gini coefficient, popular among economists, cannot be additively partitioned into within-group and between-group components.

Variance, as a measure of inequality, is faulted by economists for its lack of scale-invariance (e.g., Shorrocks, 1984). Variance in the same distribution of income in dollars, euros, yen, or any other currency is not invariant. This can be solved by care in interpreting the results, and is of less concern in demographic applications where units of, say, lifespan are less diverse than units of currency.

The interpretation of variance as a measure of inequality is helped by noting that it can be written as

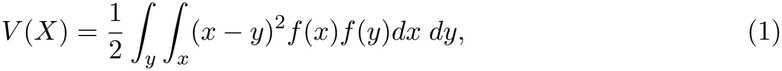

where *x* and *y* are two values drawn from the common distribution *f* (*·*). Thus the variance is one half the average squared difference between any two individuals in the population. (The Gini coefficient has the same form, but with the squared difference (*x − y*)^2^ replaced by the absolute difference |*x − y*| (Kendall and Stuart, 1969, Section 2.22).)The other useful expression for the variance is in terms of the moments of the distribution of the variable under consideration. As is well known, *V* (*X*) is the difference between the second moment and the square of the first moment,

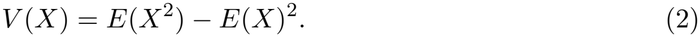

We will use this expression, with the addition of a weighting factor, in calculating lifespan inequality in heterogeneous populations. Suppose we want the variance of the entries of a vector **x**, with the entries of **x** weighted by a distribution ***ρ***. This weighted variance is

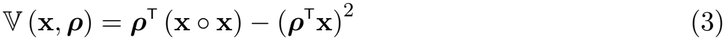

where *◦* denotes the Hadamard, or element-by-element product.

### 1.2 From one factor to multiple factors

Until recently, attempts to partition inequality into within-group and between-group components have been limited to analyzing single factors (many examples are reviewed by Caswell 2023). Even when multiple factors are addressed in a study, authors have had no choice but to analyze them one factor at a time. For example, in an admirable study, van Raalte et al. (2012) examined lifespan inequality across genders, countries, and education level. With a focus on education, they partitioned inequality into within-group and between-group components separately for each gender-country combination. Similarly, Luy et al. (2015), examined effects of sex, income, education, and work status on life expectancy, but had to examine each factor separately.

It is now possible to go beyond this limitation. In multi-factorial studies like those of van Raalte et al. (2012) and Luy et al. (2015) it is now possible to partition inequality into contributions from each factor and all combinations of two or more interacting factors (Caswell and van Daalen, 2025). For example, the variance in longevity in a population characterized by race and sex would be partitioned into *V*_race_, *V*_sex_, *V*_race*×*sex_, and *V*_stochasticity_. Caswell and van Daalen (2025) provide several examples of factorial variance partitioning, and Caswell (2026) presents a detailed four-factor analysis (sex, marital status, education, and race) based on data from Bergeron-Boucher et al. (2024).

These new methodological results apply only to factorial study designs in which all combinations of factors occur. Thus, for example, in a study treating race and sex as factors all racial categories must occur within each sex and each sex must be found in every racial category. Important as factorial studies are (and they are very important), they do not exhaust the possibilities.

### 1.3 Hierarchical study designs and data structures

In this paper, I consider the important case of studies with a hierarchical design. Such studies do not satisfy the requirement of factorial designs because some factors appear within only one category of another factor. One of these is said to be nested within the other. For the purposes of this paper I will call the groups ‘Regions,’ thinking of the regions into which the United Nations divides the countries of the world. Within each region are populations that I will call ‘Countries;’ see Figure 1.

**Figure 1:**
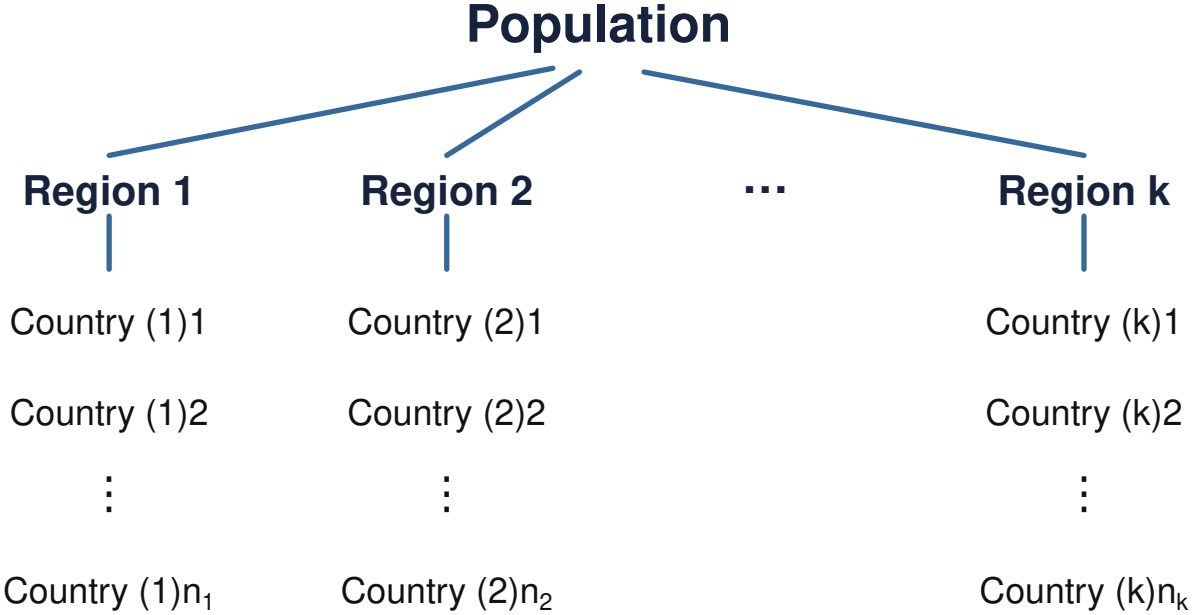
A hierarchical study design in which countries are nested within regions. Regions are numbered from 1 to *k*; countries are numbered from 1 to *n*_*i*_ in region *i* for *i* = 1, …, *k*. The parenthetical index on countries denotes the region within which the country is nested.

Countries differ in their rates, but in the absence of any further subdivision, all individuals within a country are subject to the same rates. You are free to substitute other names for the levels in the hierarchical design, **but whatever you do, do not suppose that the analysis to be presented here apply only to countries and regions**.

How important are such study designs? In fact, the largest demographic databases in existence have this structure. To cite a few examples:

1. The United Nations World Population Prospect identifies eight geographical regions based on the Sustainable Development Goals^4^ and tabulates lengthy time series of survival and fertility rates for over 200 countries distributed within these regions. The 200 countries represent demographically important factors, but each country appears within only one region.
2. The World Bank classifies countries into four income categories (high income, upper middle income, lower middle income, low income).^5^ based on inflation-adjusted gross national income per capita (GNI). Countries are nested within each of these categories; each country appears within only one income category. The Global Burden of Disease Study (e.g., GBD 2023 Disease and Injury and Risk Factor Collaborators, 2025) provides data classified, in the most recent output, into 204 countries, which are nested within 21 regions, which in turn are nested within 7 super-regions. Alternatively, countries are classified into quintiles of a Socio-Demographic Index, which integrates income per capita, average years of education, and fertility rates for women younger than 25.
3. Political classifications naturally lead to hierarchical study designs, sometimes with more than two levels. For example, life expectancy in the United States has been studied at the regional level (Vierboom, Preston, and Hendi, 2019), the state level (Montez et al., 2020), and the county level (Dwyer-Lindgren et al., 2017, 2022). Each of these levels is nested within the previous one: a county appears within only one state, a state appears within only one region.
4. In a detailed analysis of lifespan inequality, Murray et al. (2006) and Dwyer-Lindgren et al. (2024)defined first eight, and later ten, ‘Americas’ based on race, ethnicity, income, location, and racial segregation. They then analyzed and reported life expectancy in counties nested within each of these Americas.
5. In the biodemographic context, taxonomic categories are hierarchical by design, and have been ever since Linnaeus. Within the tree of life, the classification of countries within sub-regions, sub-regions within regions, and so on is replaced by species within genera, genera within families, families within orders, and so on. For example, Staerk et al. (2025) studied the evolution of sex differences in life expectancy in mammals and birds at different phylogenetic levels. The development of demographic databases for plants and animals (Salguero-Gómez et al., 2015, 2016; Species 360, 2024; Ronget et al., 2026) will make it possible to explore variance in nested sequences of phylogenetic categories.

## 2 Partitioning variance: Heterogeneous populations as nested mixtures

Accounting for variance at multiple levels requires some flexible notation. A first attempt at such is given in Table 1.

**Table 1.**
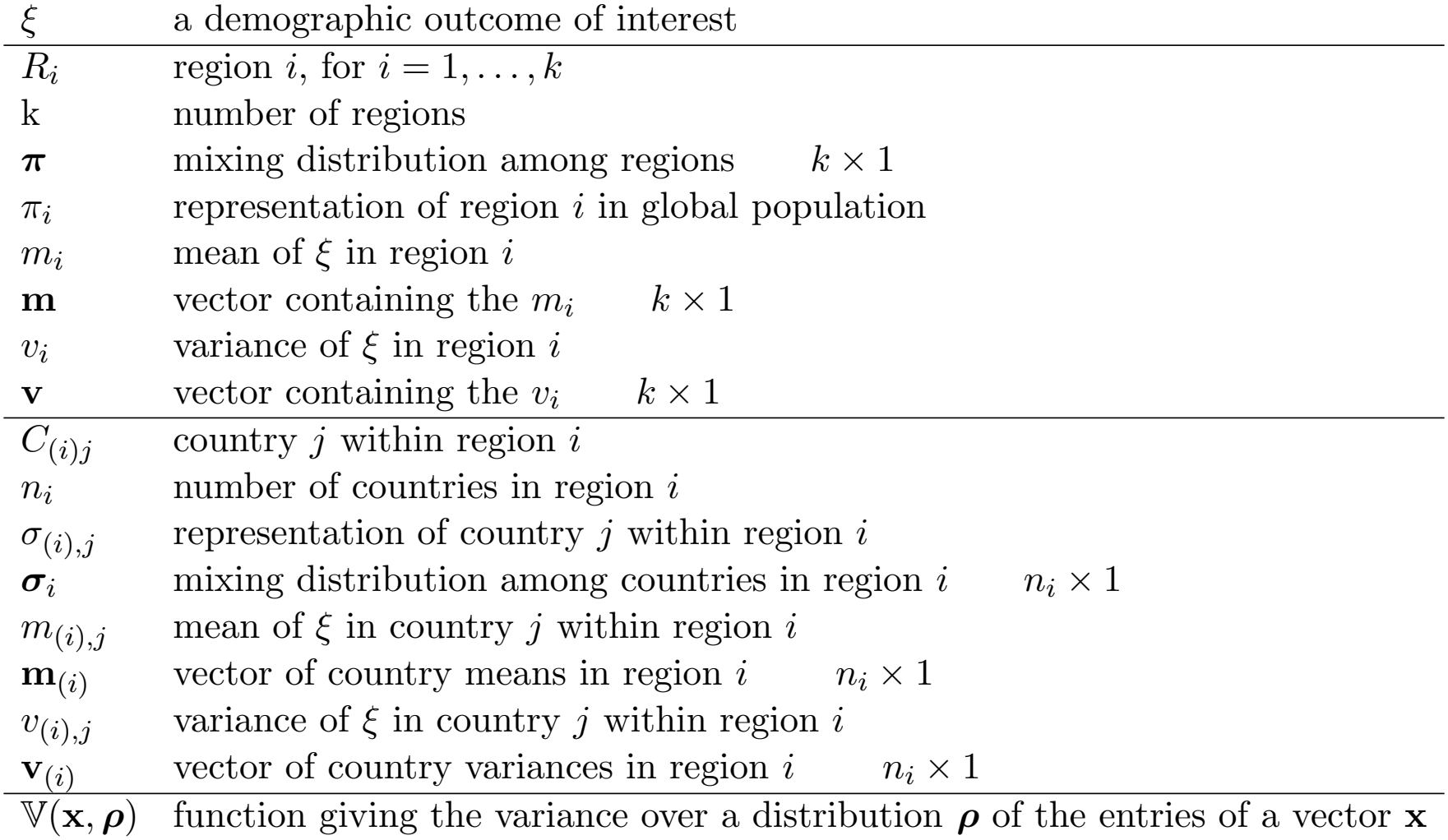
Notation for hierarchical study design. A global population is divided into regions, each of which is divided into countries.

| ξ | a demographic outcome of interest |
|---|---|
| R<sub>i</sub> | region i, for i = 1, . . . , k |
| k | number of regions |
| **π** | mixing distribution among regions k × 1 |
| π<sub>i</sub> | representation of region i in global population |
| m<sub>i</sub> | mean of ξ in region i |
| **m** | vector containing the m<sub>i</sub> k × 1 |
| v<sub>i</sub> | variance of ξ in region i |
| **v** | vector containing the v<sub>i</sub> k × 1 |
| C<sub>(i)j</sub> | country j within region i |
| n<sub>i</sub> | number of countries in region i |
| σ<sub>(i),j</sub> | representation of country j within region i |
| **σ**<sub>i</sub> | mixing distribution among countries in region i n<sub>i</sub> × 1 |
| m<sub>(i),j</sub> | mean of ξ in country j within region i |
| **m**<sub>(i)</sub> | vector of country means in region i n<sub>i</sub> × 1 |
| v<sub>(i),j</sub> | variance of ξ in country j within region i |
| **v**<sub>(i)</sub> | vector of country variances in region i n<sub>i</sub> × 1 |
| 𝕍(**x**, **ρ**) | function giving the variance over a distribution **ρ** of the entries of a vector **x** |

The key to partioning inequality in hierarchical study designs is to treat the distribution of lifespan in the heterogeneous population as a mixture distribution. A heterogeneous population is a mixture made up of groups of individuals that differ in the demographic rates to which they are subject (Caswell, 2023). Figure 2 shows an example in which the population is divided into *k* ‘regions,’ each characterized by a mean and variance of some demographic outcome *ξ*, and with a representation *π*_*i*_ in the population.

**Figure 2:**
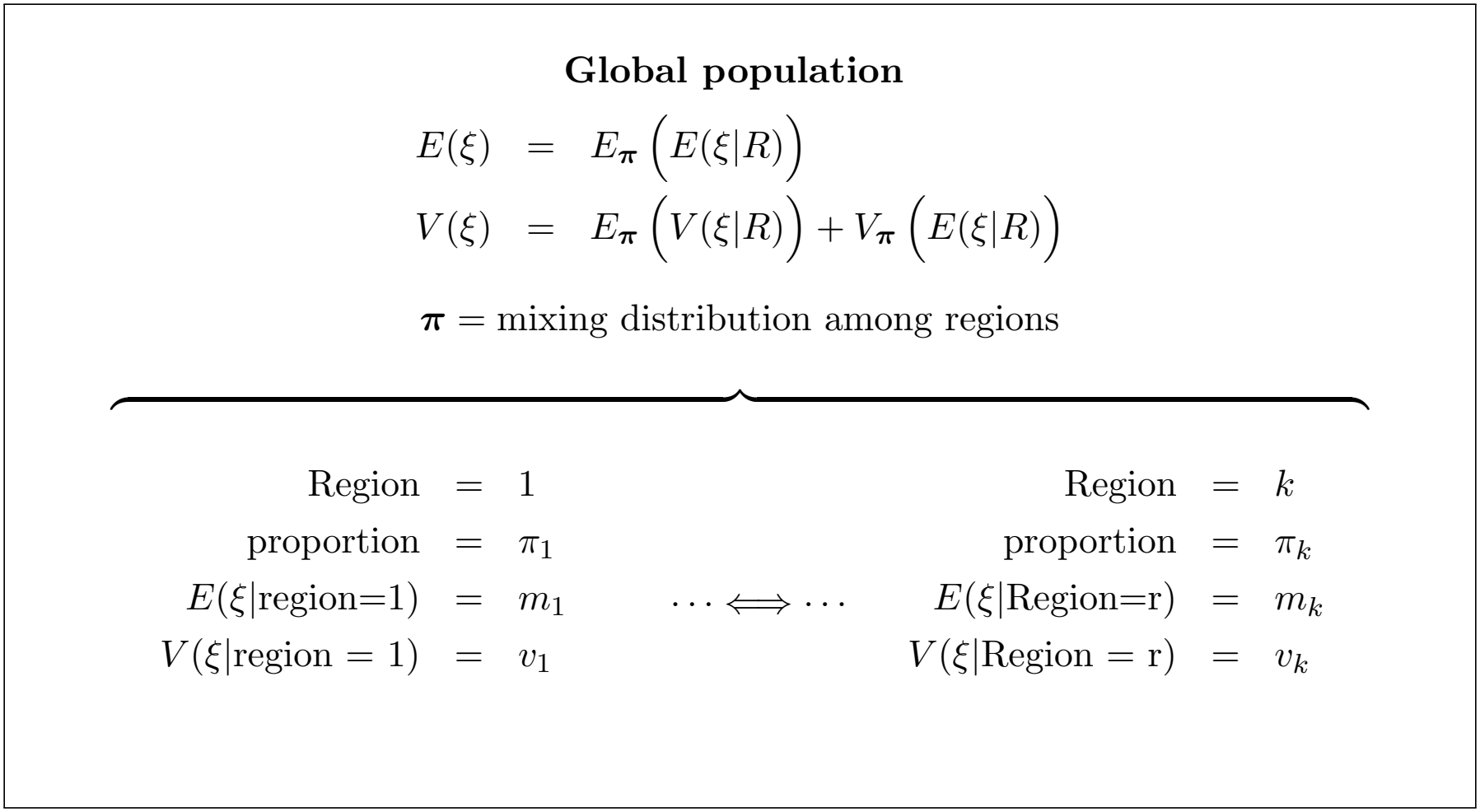
A heterogeneous population composed of a mixture of regions, from *R* = 1 to *R* = *k*, with mixing distribution ***π***. Modified from Caswell and van Daalen (2025) under the terms of a CC-BY license.

Recall that a variate *X* has a (finite) mixture distribution if its probability distribution *h*(*x*) can be written as a linear combination of distributions *f*_1_(*x*), …, *f*_*k*_(*x*),

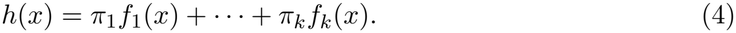

The coefficients *π*_*i*_ are referred to as the mixing distribution. The moments of a mixture distribution are weighted means of the moments of the *f*_*i*_, and the variance of the mixture distribution can be calculated, as we will see, from the means and variances of the *f*_*i*_.

The study design in Figure 2 contains only one level of heterogeneity: that among regions. But each region is made up of a set of countries. Figure 3 focuses in on Region 1, showing that it is a mixture of *n*_1_ countries, each with a mean and variance of *ξ* distributed according to a mixing distribution ***σ***_1_.

**Figure 3:**
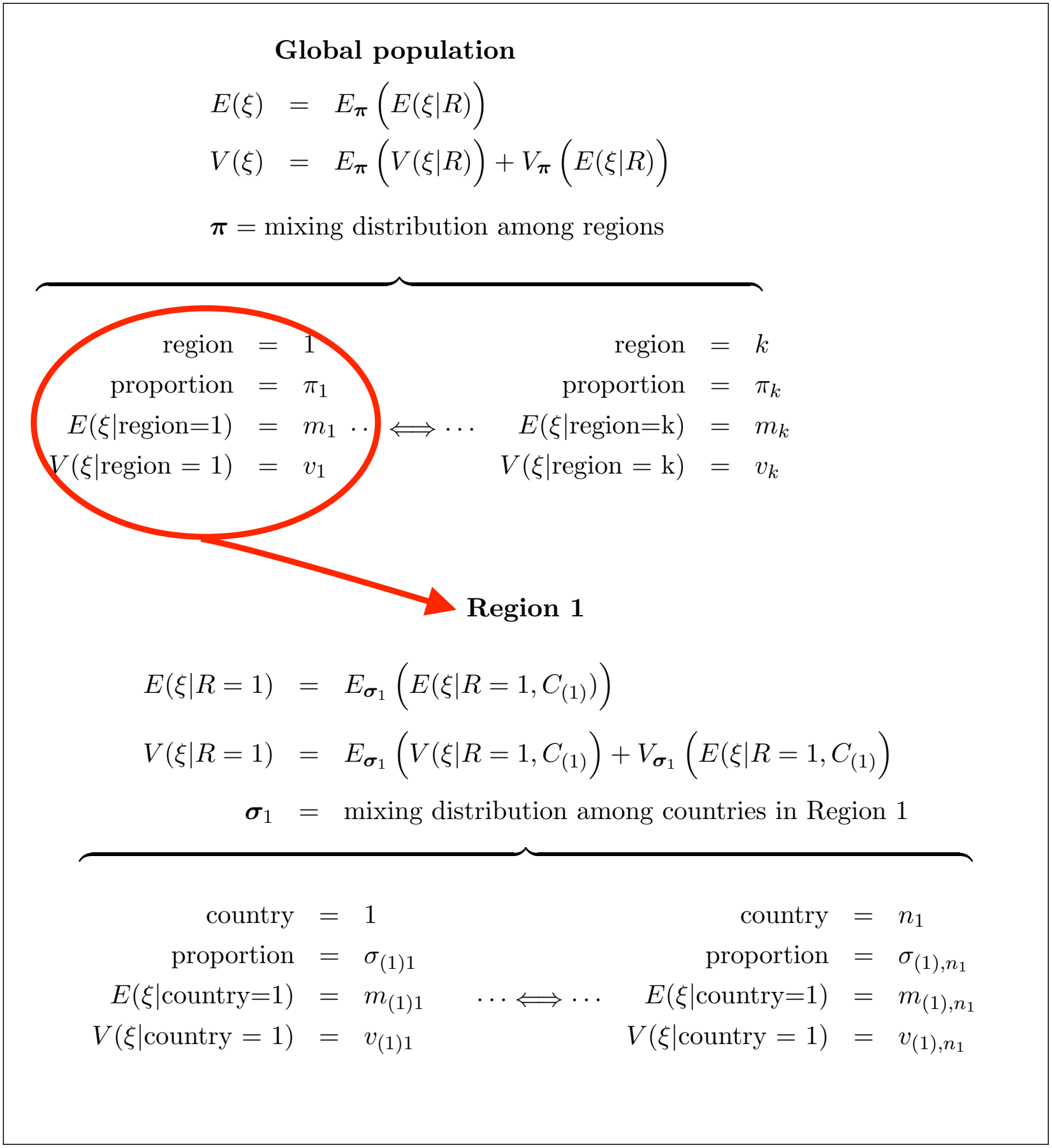
A closer look at Region 1 of the multi-region structure of Figure 2 Each region of the global population is itself a mixture of countries. Region *i* contains *n*_*i*_ countries, with a mixing distribution ***σ***_*i*_.

### 2.1 Means within regions and within countries

As shown in Figure 2, the overall population is a mixture of *r* regions. Each region is characterized by the mean and variance of some variable *ξ*. The mixture of regions that make up the population is defined by a mixing distribution ***π***, where the entries of ***π*** are non-negative and sum to 1.

A closer look at the structure of Figure 2, as shown in Figure 3, recognizes that each Region *i* is composed of some number *n*_*i*_ of countries, each with its mean and variance, and distributed according to a mixing distribution ***σ***_*i*_.

The mean of *ξ* in the overall population is

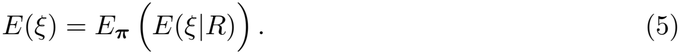

The subscript on *E*_***π***_ denotes the probability distribution over which the expectation is calculated, which is the distribution of the random variable *R* on which *E*(*ξ*|*R*) is conditioned; see Appendix A. Thus,

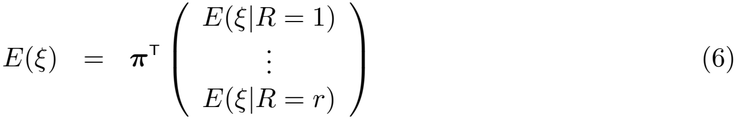

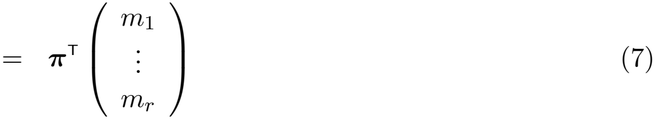

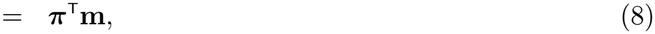

where **m** is a vector, of dimension *r ×* 1, whose entries are the means in each of the regions.

The mean *m*_*i*_ in region *i* is itself an expectation, taken over a set of country means,

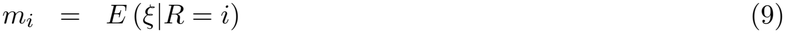

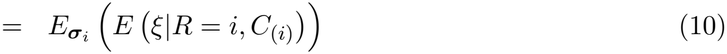

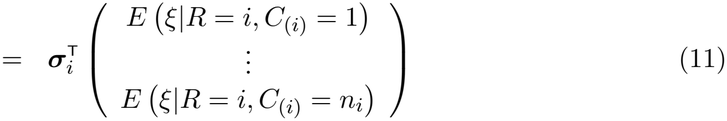

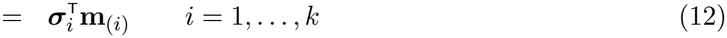

Where *C*_(*i*)_ is the random variable denoting countries within region *i* and **m**_(*i*)_ is the vector of country means within region *i*. Thus, substituting Equation (12) into (8) we get the unconditional mean of *ξ* over the whole global population as

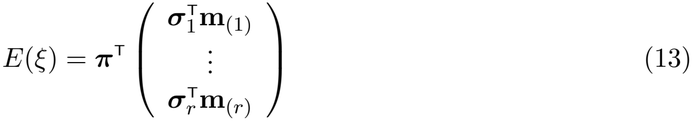

### 2.2 Variances within regions and countries

The variance of *ξ* in the global population has two components, which are is given by the Law of Total Variance applied to the regions within the population:

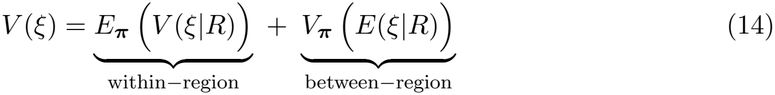

The within-region variance is the expectation, over the mixing distribution ***π***, of the region-specific variances. The between-region variance is the variance, over the mixing distribution, of the region-specific means. Together, these two components account for all the variance in *ξ* at the overall population level.

Let us consider the two terms in order. The within-region variance is

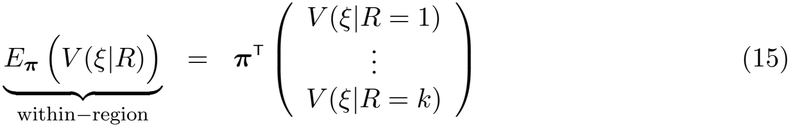

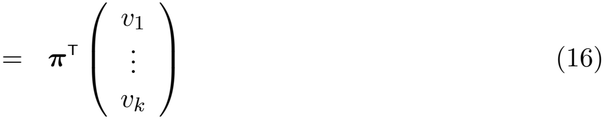

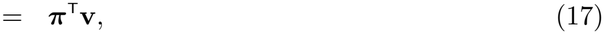

where *v*_*i*_ is the variance within region *i* and **v** is a vector, of dimension *r ×* 1, containing the *v*_*i*_.

In the hierarchical study design, the variance within region *i* also has two components: one within, and one between, the countries that make up region *i*.

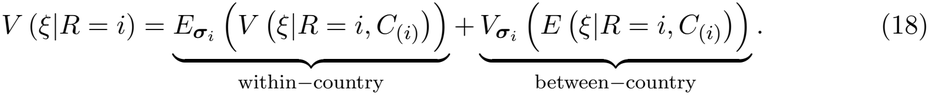

The within-country variance in equation (18) is the mean, over the mixing distribution ***σ***_*i*_, of the variances within countries in region *i* and is given by

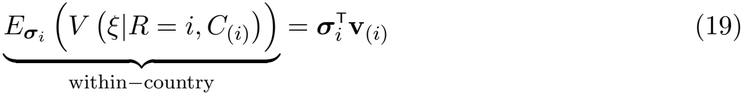

The between-country variance in Equation (18) is the variance, over the distribution ***σ***_*i*_, among the means of the countries in region *i*. It is obtained by applying Equation (3) to those means,

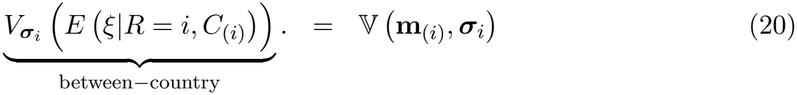

Returning to the between-region variance in Equation (14), it is obtained by applying equation (3) to the means of *ξ* conditional on regions:

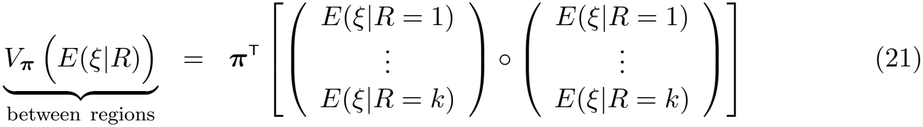

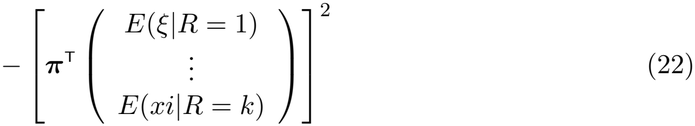

which, thankfully, can be written more compactly as

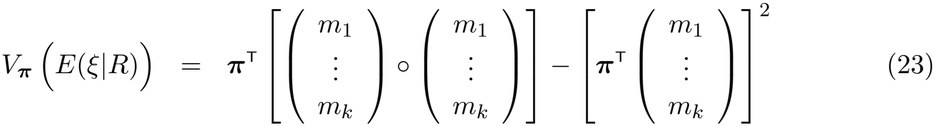

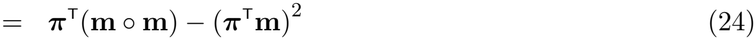

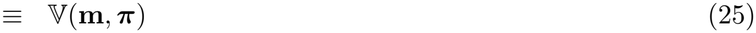

### 2.3 Combining levels: From countries to regions to the population

The result of all this (tedious but necessary) algebra is a set of expressions for the means and variances of the outcome *ξ* at all levels of the hierarchical design. The means within the regions are given in Equation (12). These means are combined in Equation (8) to give the overall population mean.

The global variance is more complicated. Of course it is.

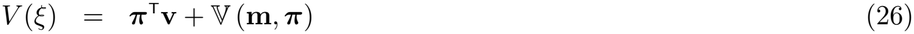

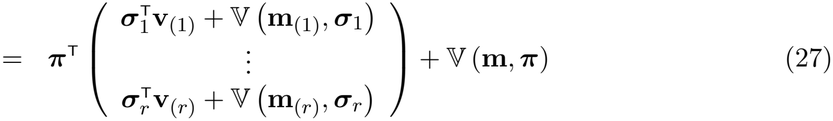

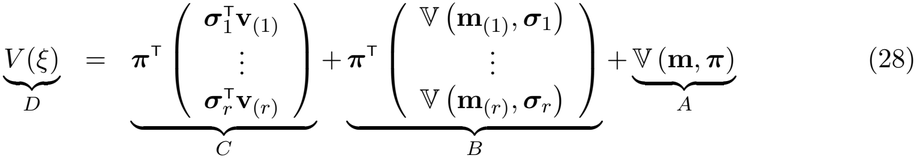

We have now partitioned the variance into three components, as identified in Equation (28):

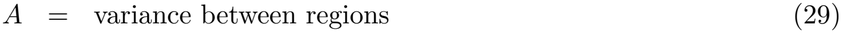

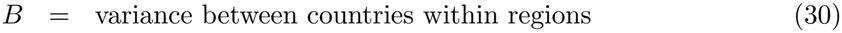

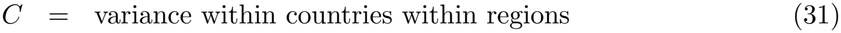

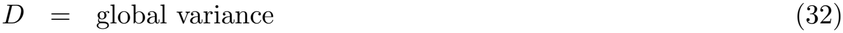

Working from right to left, the first component (*A*) is the contribution of heterogeneity among the regions making up the population, as shown in Figure 2. The second component (*B*) is the contribution of heterogeneity among the countries making up the regions, as shown in Figure 3. The final component (*C*) is the contribution of stochasticity within countries. These components can be tabulated as in Table 2.

**Table 2.** Variance partitioning for a demographic outcome *ξ* between regions, between countries within regions, and due to stochasticity within countries.

| source | variance |
|---|---|
| between regions | *V*<sub>*R*</sub> |
| between countries within regions | *V*<sub>(*R*)*C*</sub> |
| within countries (stochasticity) | *V*<sub>*W*</sub> |
| total | *V* |

**Table 3.** The definitions of low, lower-middle, upper-middle, and high income regions, based on value of GNI per capita.

| World Bank GNI intervals 2024–2025 |  |
|---|---|
| low income | ≤ \$1, 145 |
| lower-middle income | \$1,146–\$4,515 |
| upper-middle income | \$4,516–\$14,05 |
| high income | > \$14, 005 |

## 3 An example: Global inequality in healthy longevity

Zarulli and Caswell (2024) presented a global stochastic analysis of inequality in healthy longevity. That analysis was based on data from the Global Burden of Disease (GBD) study (GBD 2021 Diseases and Injuries Collaborators, 2024).

The GBD study computed years lived with disability, years of life lost, disability-adjusted life years, and healthy life expectancy (HALE) for 371 diseases and injuries IN nearly 200 countries and territories, over the years 2010 to 2021. Like any measure of life expectancy, HALE is an expected value or mean of lifespan. Zarulli and Caswell (2024) applied a Markov chain with rewards model (van Daalen and Caswell, 2017; Caswell and Zarulli, 2018) to the GBD data obtain the variance of healthy longevity. The variance, which describes the differences among individuals implied by the age-schedule of mortality and disability, reflects the stochastic nature of the demographic rates. It measures inequality of this important demographic outcome among individuals subjected to a specified set of vital rates. Zarulli and Caswell reported time trends of both HALE and SDHL for different ages and countries.

As an example of a hierarchical variance partition, in this paper I use the GBD data to examine inequality in healthy lifespan in a global perspective. GBD includes two hierarchical levels of heterogeneity in their analysis. The first is differences among regions defined by the categories of Gross National Income (GNI) per capita as defined by the World Bank: The second is due to the differences among the countries included in those income groups (31, 45, 59, and 63 countries in the four regions, respectively).

### 3.1 Mixing distributions for the global model

The variance components were calculated using equation (28). Flat mixing distributions, for both ***σ***_*i*_ and ***π***, were used to give equal representation to each region and each country within each region. A proportional, population-weighted mixing distribution was created using population sizes for each country, as reported in the database created by Jorda, Niño-Zarazúa, and Tejería-Martínez (2024), based on the 2022 revision of the United Nations World Population Prospects.

### 3.2 Results: healthy longevity in a global perspective

When applied to the data of Zarulli and Caswell (2024), the analysis provides the results on the mean and variance of lifetime healthy longevity from birth, from all starting ages, over a period of 30 years, for females and for males, and subject to both flat and population-weighting distributions. In this section we will explore some of these results, focusing mainly on females with population-weighted mixing distributions. The corresponding results for males and for the flat mixing distribution are given in the Supplementary Material.

#### 3.2.1 Healthy life expectancy

Table 4 and Figure 4 show mean healthy longevity (i.e., healthy life expectancy) for females and males, in 1990 and 2019. Differences among income regions are substantial. Healthy life expectancy in the high-income region exceeds that in the low-income region by 22 years for women in 1990 and by 13 years in 2019. The corresponding differences for men are 20 years and 13 years. Healthy life expectancy is, as usual, higher for women than for men, by 1–4 years. Between-year differences are also apparent, especially in the low-income region. Healthy longevity increased by 12 years for both females and males between 1990 and 2019.

**Table 4.** The mean healthy longevity (i.e., health expectancy) in years, for men and women, in each of the four regions defined by the World Bank income classes, in 1990 and in 2019.

|  |  | Region |  |  |  |
|---|---|---|---|---|---|
|  |  | low | lower-mid | upper-mid | high |
| 1990 | female | 45.3 | 52.3 | 61.5 | 67.6 |
|  | male | 43.8 | 51.6 | 58.6 | 63.8 |
| 2019 | female | 57.6 | 61.0 | 69.1 | 71.0 |
|  | male | 55.5 | 59.7 | 65.8 | 68.9 |

**Figure 4:**
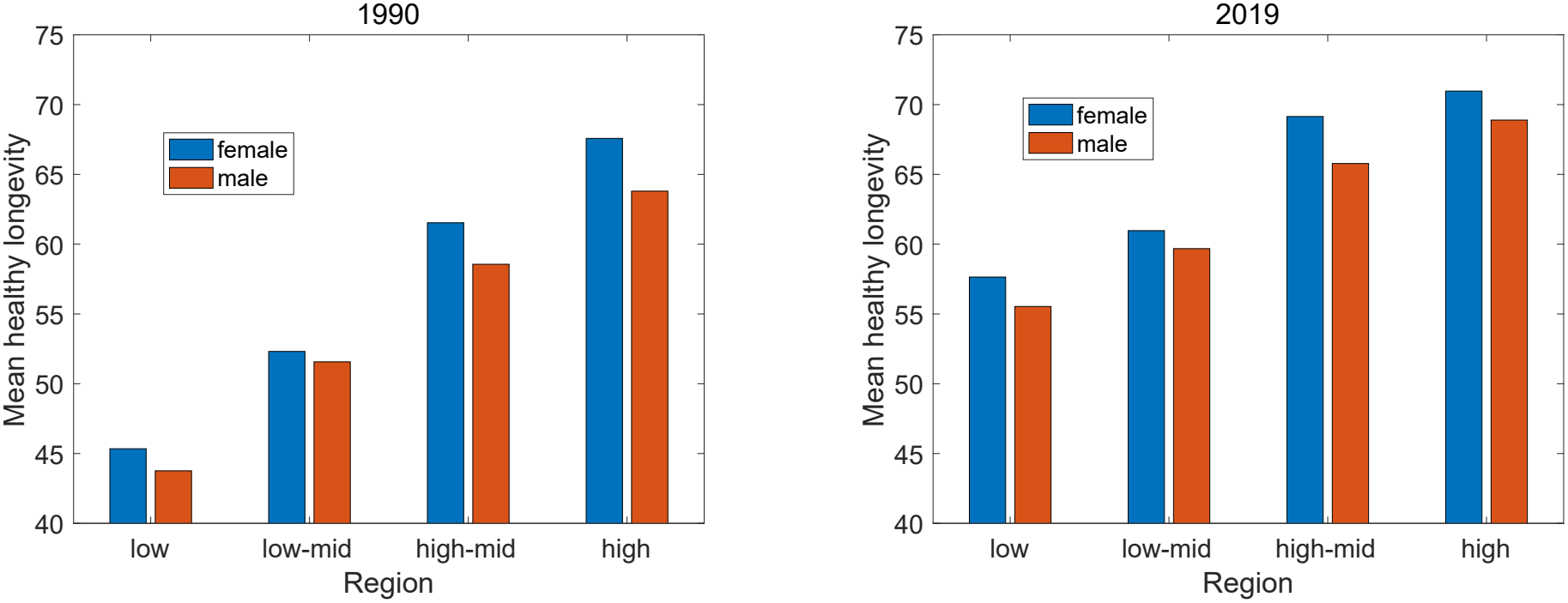
Mean remaining healthy longevity for males and females, across four income regions, in 1990 and 2019. Population-weighted mixing distribution.

#### 3.2.2 Variance partitioning

Knowing the mean healthy longevity tells nothing about the differences, or inequalities, between individuals. Table 5 begins our exploration of the contributions from differences between regions, differences between countries within regions, and stochastic variation within countries to the variance in healthy longevity. Results are shown for healthy longevity measured from birth and for remaining healthy longevity from age 60, using mortality rates for 1990 and 2019 and both flat and population-weighted mixing distributions.

**Table 5.**
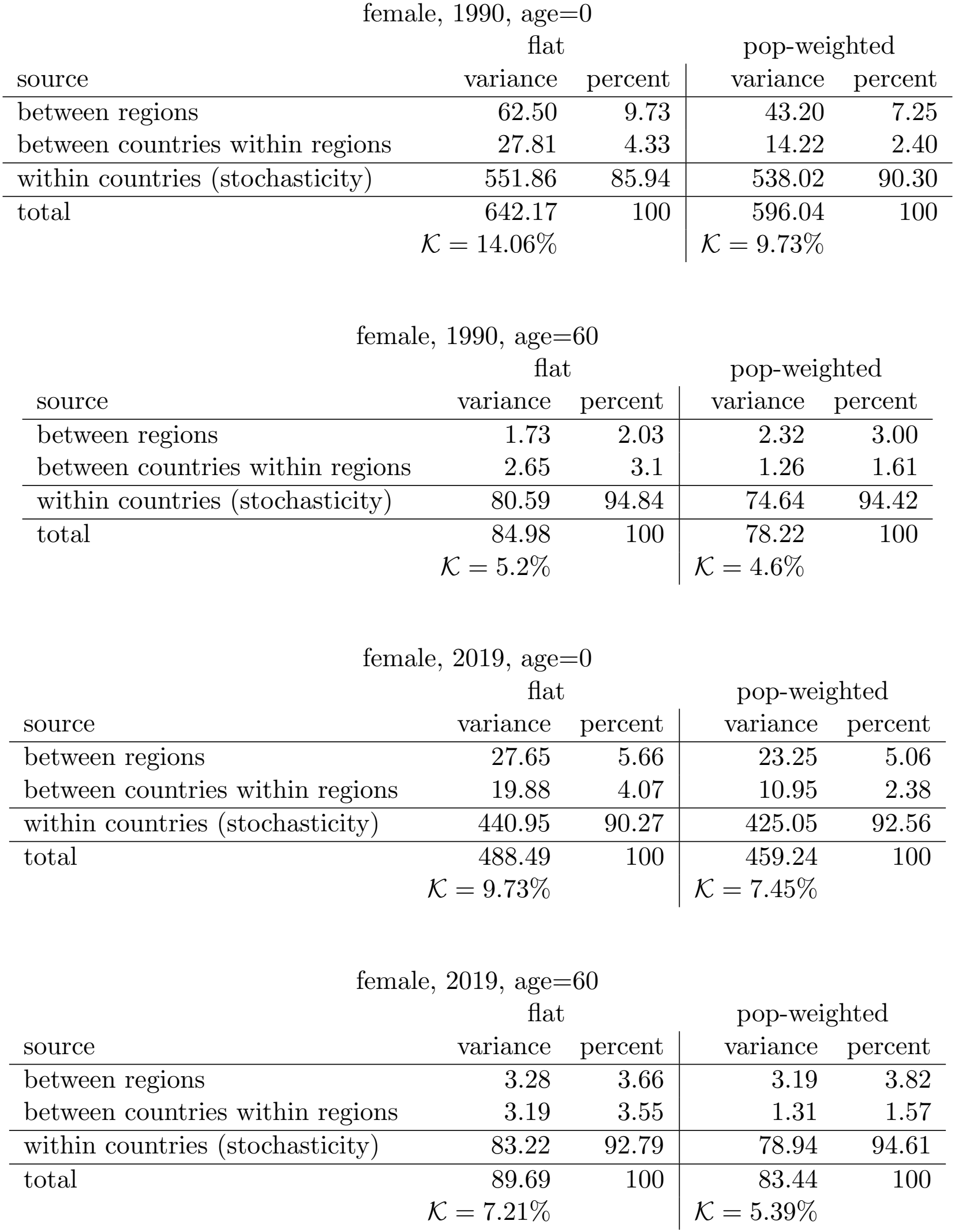
Partition of the variance in remaining healthy longevity, at birth and at age 60, into contributions due to regions, to countries within regions, and due to individual stochasticity within countries. Rates of the year 1990 and 2019. Mixing distributions flat and proportional to population sizes of the countries and regions.

| female, 1990, age=0 |  |  |  |  |
|---|---|---|---|---|
|  | flat |  | pop-weighted |  |
| source | variance | percent | variance | percent |
| between regions | 62.50 | 9.73 | 43.20 | 7.25 |
| between countries within regions | 27.81 | 4.33 | 14.22 | 2.40 |
| within countries (stochasticity) | 551.86 | 85.94 | 538.02 | 90.30 |
| total | 642.17 | 100 | 596.04 | 100 |
|  | 𝒦 = 14.06% |  | 𝒦 = 9.73% |  |
| female, 1990, age=60 |  |  |  |  |
|  | flat |  | pop-weighted |  |
| source | variance | percent | variance | percent |
| between regions | 1.73 | 2.03 | 2.32 | 3.00 |
| between countries within regions | 2.65 | 3.1 | 1.26 | 1.61 |
| within countries (stochasticity) | 80.59 | 94.84 | 74.64 | 94.42 |
| total | 84.98 | 100 | 78.22 | 100 |
|  | 𝒦 = 5.2% |  | 𝒦 = 4.6% |  |
| female, 2019, age=0 |  |  |  |  |
|  | flat |  | pop-weighted |  |
| source | variance | percent | variance | percent |
| between regions | 27.65 | 5.66 | 23.25 | 5.06 |
| between countries within regions | 19.88 | 4.07 | 10.95 | 2.38 |
| within countries (stochasticity) | 440.95 | 90.27 | 425.05 | 92.56 |
| total | 488.49 | 100 | 459.24 | 100 |
|  | 𝒦 = 9.73% |  | 𝒦 = 7.45% |  |
| female, 2019, age=60 |  |  |  |  |
|  | flat |  | pop-weighted |  |
| source | variance | percent | variance | percent |
| between regions | 3.28 | 3.66 | 3.19 | 3.82 |
| between countries within regions | 3.19 | 3.55 | 1.31 | 1.57 |
| within countries (stochasticity) | 83.22 | 92.79 | 78.94 | 94.61 |
| total | 89.69 | 100 | 83.44 | 100 |
|  | 𝒦 = 7.21% |  | 𝒦 = 5.39% |  |

Across all scenarios, the dominant source of variation is stochasticity within countries. Depending on the year, age, and mixing distribution, between-group heterogeneity (both between regions and between countries) accounts for only about 5% to 14% of the total variance, while the remaining 86% to 95% is attributable to stochastic variation within countries. Thus, although systematic differences among regions and countries are evident, most inequality in healthy longevity reflects stochastic variation among individuals subject to the same country-specific mortality and disability rates.

The relative importance of heterogeneity changes with age. Healthy longevity measured from birth exhibits substantially greater overall variance than remaining healthy longevity at age 60. Under the flat mixing distribution, the total variance declines from 642.2 to 85.0 in 1990 and from 488.5 to 89.7 in 2019. This reduction is accompanied by a decline in the proportion of variance attributable to heterogeneity, from 14.1% to 5.2% in 1990 and from 9.7% to 7.2% in 2019. Remaining healthy longevity at age 60 is therefore even more strongly dominated by stochastic variation than healthy longevity measured from birth.

Population weighting reduces the contribution of between-group heterogeneity in all four comparisons in Table 5. The reduction is especially pronounced for differences among countries within regions. For example, at birth in 2019, the contribution of between-country variation declines from 4.1% under flat mixing to 2.4% under population weighting, while the regional contribution changes from 5.7% to 5.1%. The variance ratio K declines from 9.7% to 7.5%. Despite these differences between mixing distributions, stochasticity remains by far the dominant source of variation.

Table 5 thus provides snapshots of the variance decomposition at two ages and in two years. The following sections examine changes in these components continuously across age and calendar time.

#### 3.2.3 Changes in variance components over age

Figure 5 shows the components of variance in remaining healthy longevity as a function of age, in 1990 and in 2019, for females under the population-weighted mixing distribution.^6^ The overall variance declines consistently from birth to age 85, at which point the life table is truncated. Although both stochastic and between-group components also decline, their relative rates of decline differ, producing a nonlinear age pattern in the variance ratio *K*, which declines most sharply at younger and older ages, with comparatively little change through much of adulthood. Stochasticity within countries dominates the variance at all ages.

**Figure 5:**
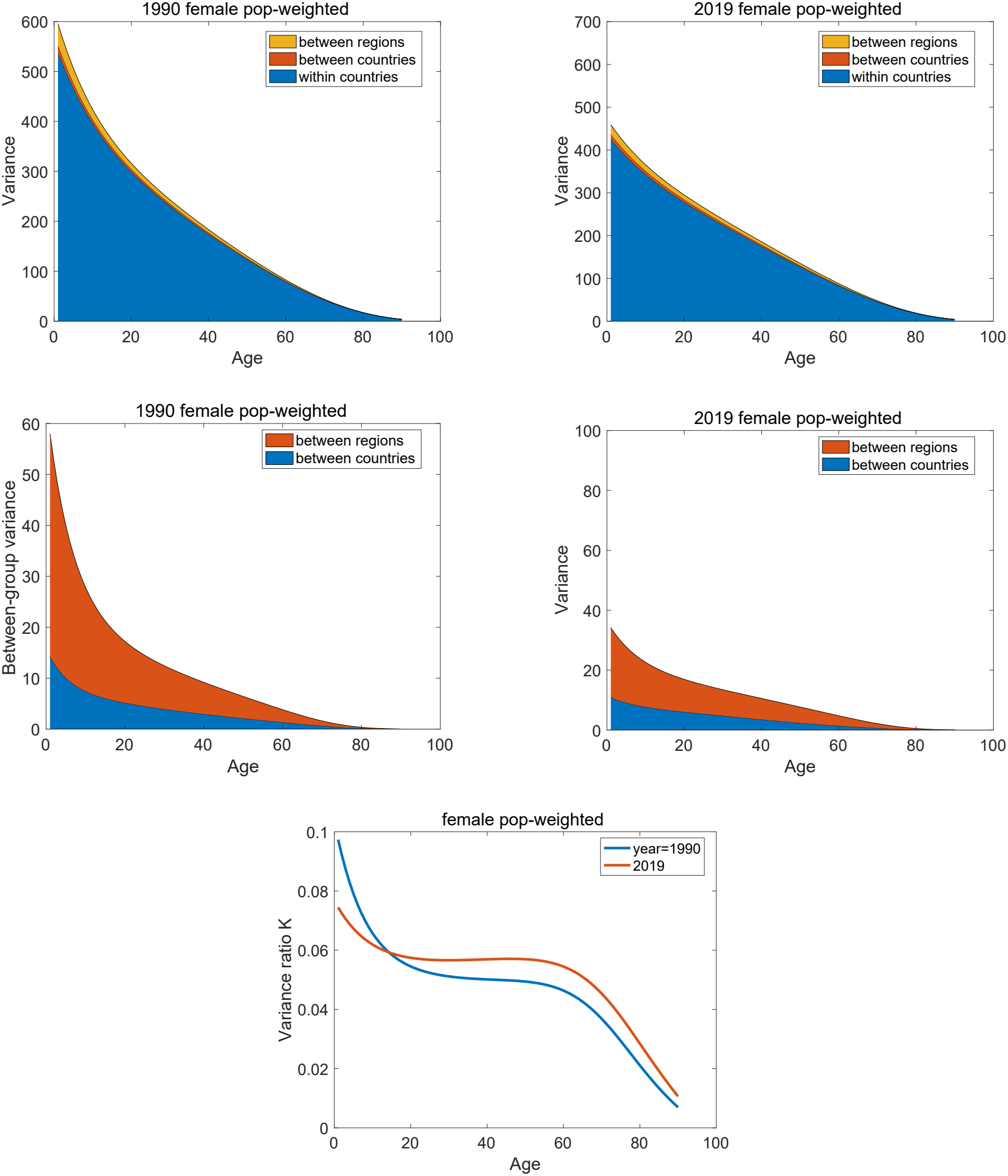
Variance components and the variance ratio as functions of age, for females with the population-weighted mixing distribution, in 1990 and 2019.

The changes in variance components with age are qualitatively similar for males, and for flat as contrasted with population-weighted mixing distributions (Appendix C).

#### 3.2.4 Changes in variance components over time

Figure 6 shows the temporal trends in the components of variance of healthy longevity for females of selected ages, using a population-weighted mixing distribution. Temporal trends in total variance differ markedly by age. From 1990 to 2019, variance measured at birth declines substantially. Variance at age 20 declines and variance at age 65 increases slightly, with some evidence of a peak around 2010.

**Figure 6:**
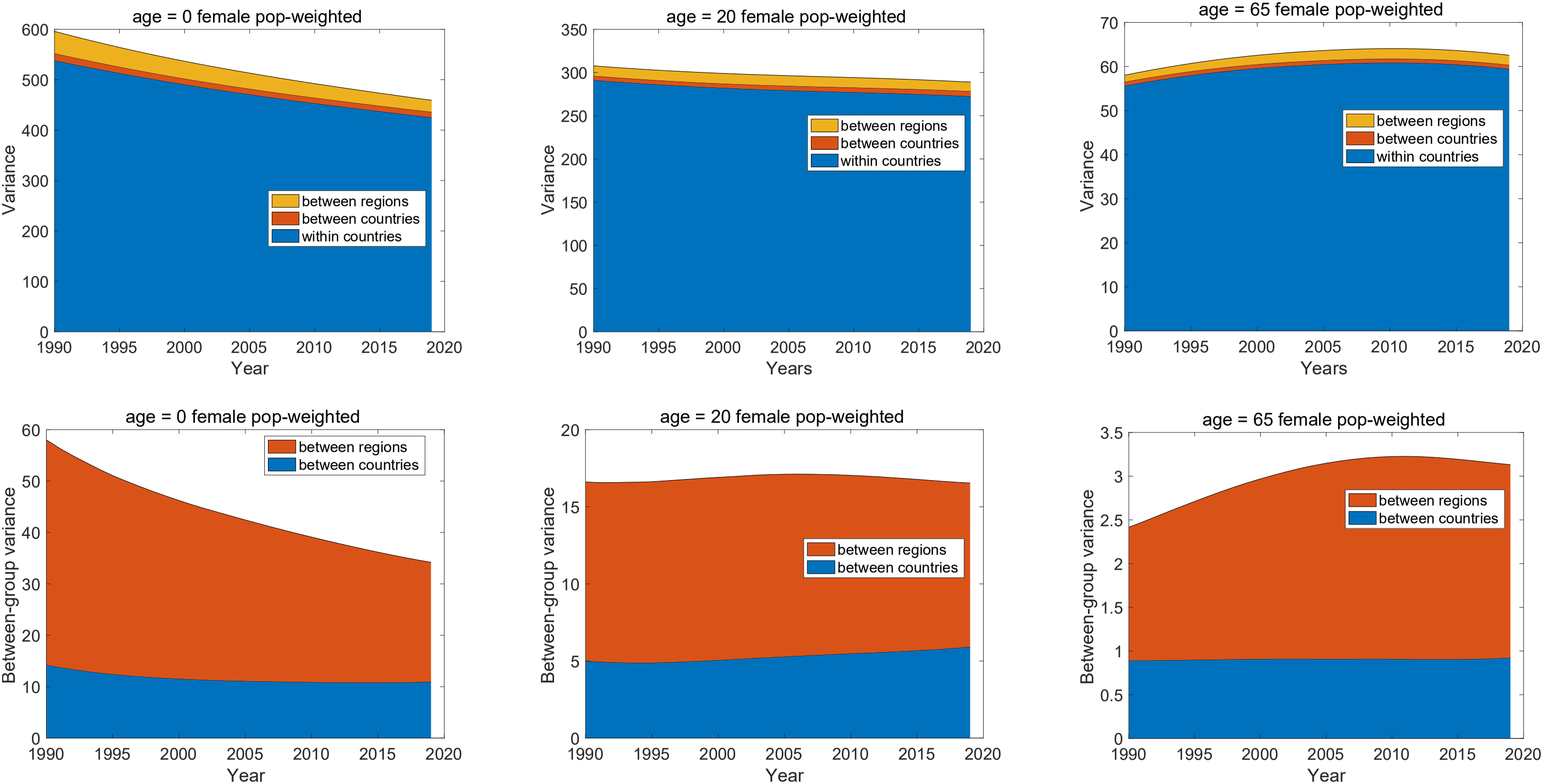
variance components for remaining female healthy longevity as a function of time, for selected ages (0, 20, 65). Note different ordinate scales.

Regional differences generally make a larger contribution to between-group variance than differences among countries within regions, although the magnitude of this difference varies with age and time. The temporal trend in K also depends on age (Figure 7). At ages 0 and 20, K declines between 1990 and 2019, whereas at age 65 it increases. Even at age 65, however, between-group heterogeneity remains small, accounting for no more than about 6% of total variance.

**Figure 7:**
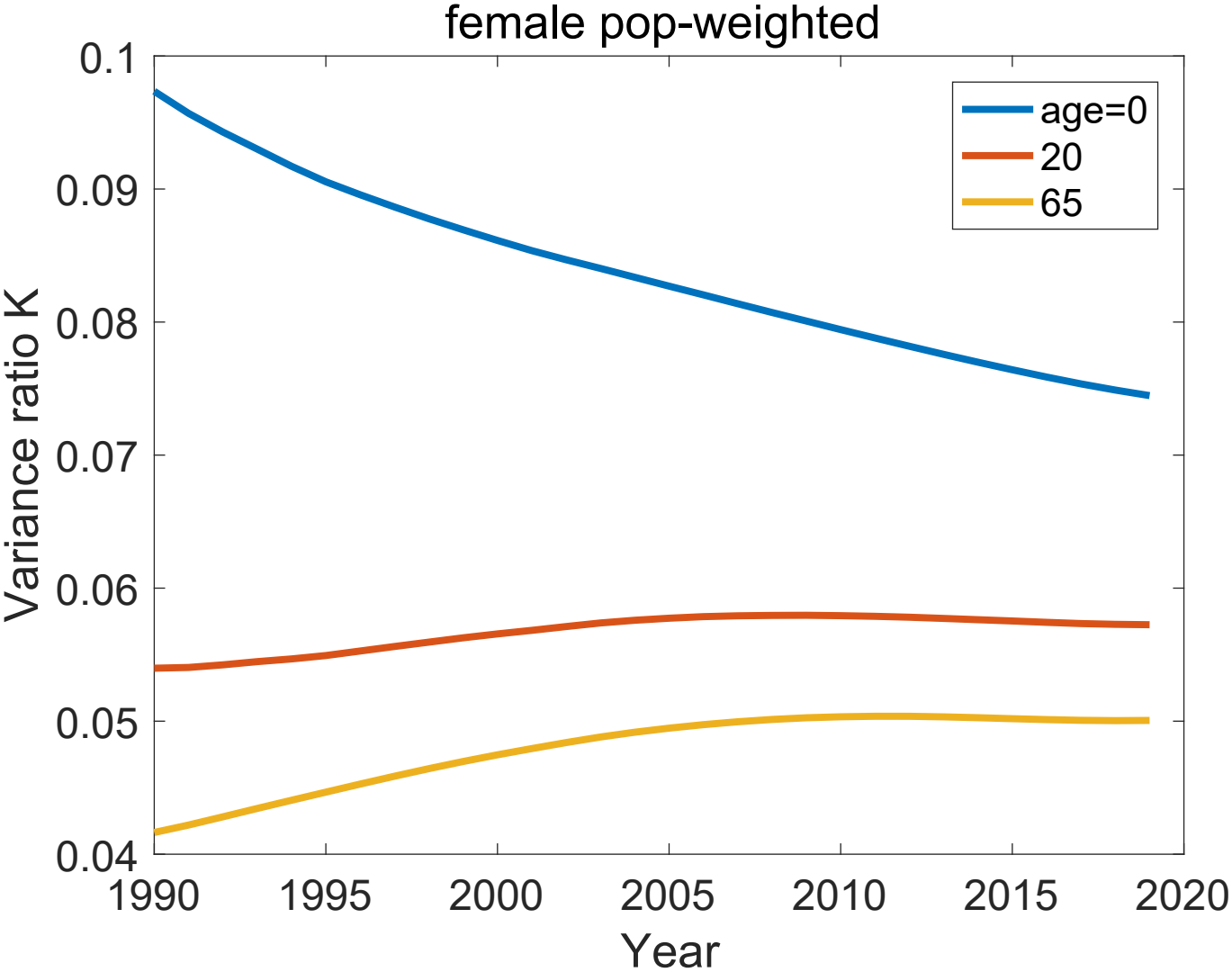
The variance ratio *K* for remaining female healthy longevity, as a function of time, for selected ages (0, 20, 65).

## 4 Discussion

### 4.1 A new methodology for hierarchical study designs

Hierarchical study designs are broadly applicable in demography. The partition of variance in these designs has been introduced here in terms of a two-level design, in which populations are divided into categories (here, regions) which in turn are divided into sub-categories (here, countries). Each level of the hierarchy contributes a within-group and a between-group component of variance. The steps in the analysis are given in Appendix B.

This two-level study design can be extended in several directions. For example, a study may include multiple nested levels (e.g., regions, countries within regions, states within countries). Or a study might combine hierarchical and factorial structures. The top level in the example here was that of World Bank income regions. Another factor (e.g., sex) could be included at the top level of the design. Inequality at the top level would then include variance components due to region, to sex, and the region*×*sex interaction. Within each region-sex combination, we would find variance between countries and stochasticity within countries.

The hierarchical design is not restricted to regions and countries. I have used that terminology only because it describes data structures like listed in Section the U.N. WPP, and is more intuitive than trying to keep track of “groups,” “sub-groups,” “sub-sub-groups” and so on.

In some cases the hierarchical structure may be ambiguous. Consider a study with sexes and countries as factors. This seems like a factorial design, in which both males and females appear in every country. But on the other hand, the experience of women and men may differ dramatically between countries, due to social, religious, and political systems. A study design that treats gender as a factor nested within, rather than cross-classified with, countries would be a way to approach this. Instead of having male and female within every country, this design would define “male and female in Country 1”, “male and female in Country 2,” and so on, as separate categories.

The partitioning of inequality is not limited to longevity or to healthy longevity. The method presented here can be applied to any demographic outcome for which means and variances can be calculated. This includes lifetime reproductive output (e.g., van Daalen and Caswell, 2017; Snyder and Ellner, 2018; Tuljapurkar et al., 2020), lifetime overlap with kin (Caswell and de Vries, 2025), and occupancy of health states in incidence-based health models (Caswell and van Daalen, 2021).

#### 4.1.1 The mixing distribution

In factorial study designs it is only possible to partition variance into main effects and interactions if the mixing distribution is either flat (all factor combinations receiving equal weight) or a rank-one distribution (all the marginal distributions are proportional). In the two-level hierarchical design considered here, both the regional mixing distribution and the country distribution within each region are one-factor designs. Thus the analysis imposes no restrictions on the mixing distributions ***π*** and ***σ***_*i*_. More complex hierarchical studies with factorial designs at the top level would be restricted to flat or rank-one distributions.

#### 4.1.2 What about interactions?

There are no interactions between factors that appear in different levels (countries and regions in our case) in a hierarchical design. The reason is not hard to see. In a factorial design with factors A and B, an A*×*B interaction implies that the effect of factor A differs depending on the level of factor B. For examples, see Caswell and van Daalen (2025); Caswell (2026) In a hierarchical analysis of regions and countries, a country appears in only one region, so it is impossible for its effect to differ from region to region. To state the case more generally, interactions cannot appear between factors that appear at two different levels in the hierarchical design.

#### 4.1.3 Is this more than just ANOVA?

The variance decomposition in Table 5 resembles a classical nested Analysis of Variance (ANOVA), but the two differ fundamentally in their data requirements. The statistical model for ANOVA begins with a vector of individual observations, written as a linear combination of variables representing factors (e.g., Gill, 1978; Kirk, 1982). Those individual observations provide the sums of squares that the model minimizes to estimate variances. Our situation is different. The life tables from which means and variances of longevity are calculated contain no individual observations of longevity. Thus it is impossible to develop an ANOVA model from life table data.

When individual data *are* available, then models based on those data can explore lifespan inequality using linear (and nonlinear) models. Breen and Seltzer (2026) do so using a painstakingly assembled data set on individual longevity and a machine learning approach to fit a model including 32 factors, including detailed information on education, race, occupation, household structure, and movements. They find statistically significant relationships between some of the factors and longevity, but those relationships explain only 1.3% of the variance; the remainder being attributed to stochasticity.

### 4.2 A global analysis of variance in healthy longevity

The dominant source of inequality in healthy longevity in this global analysis is stochastic variation within countries, which accounts for at least 85% - 95% of the overall inequality, depending on the year and the age and sex of the individuals. Stochasticity maintains its importance in the face of an enormous global range of socio-economic conditions.

In 2024-2025 the countries with the lowest GNI values were Burundi ($240) and Afghanistan ($390). The highest GNI values included Bermuda ($139,370) and Switzerland ($110,330). The difference in quality of life and health supported by these economies is undeniable, and the GNI values capture at least this aspect of economic conditions on the global scale.

Of course, countries within the same income category can differ substantially in their health systems and other social and institutional determinants of health. Such differences contribute to the between-country variance, but even after allowing for them, stochastic variation within countries remains the dominant component of variance.

This dominance of stochastic variation should not be interpreted to mean that socioeconomic or institutional differences are unimportant. Such differences can strongly affect the mean level of healthy longevity even when they account for a relatively small fraction of its individual-level variance. And they reflect differences that can be impacted, for good or ill, by policies and politics.

The results reported here are an opportunity to compare the flat and the population-weighted mixing distributions. The major difference between the two is the weight that they assign to each region or country. The flat distribution allows each country to contribute equally to each region, and each region to contribute equally to the population. This maximizes the information about the effects of ‘treatments’ (countries and regions) on *ξ* (see Caswell 2026 for discussion of multifactor studies and experiments).

Population-weighted mixing distributions treat large countries as more important than small ones, which may or may not be desirable. For example, both China (population 1.4 billion) and the Marshall Islands (population 35,000) fall in the Upper-Middle income group. The flat distribution recognizes both as countries and lets them both contribute as such. The population-weighted distribution gives China more than 40,000 times the influence of the Marshall Islands. The many differences between China and the Marshall Islands would be completely obscured by this mixing distribution, but if the question at hand is about the overall population of the upper-middle income region, it would be an appropriate distribution. Different questions, different mixing distributions.

Because the population-weighted mixing distribution concentrates influences on variance into a smaller number of countries and regions, it reduces the between-group variance and the variance ratio *K*. (Table 5). Qualitatively, however, the patterns remain: stochasticity accounts for 85%–95% of the variance, variance declines with age and, changes over time (Figures 5–7).

This global data set spans the range of economic conditions and of country characteristics. The economic conditions make a sizable impact on mean healthy longevity (Table 4 and Figure 4). Even so, only a small fraction of the inequality in healthy longevity is accounted for by these differences. That does not make them unimportant, both to demographic research and to the people living under these conditions.

## Data Availability

Sources of the data in the published literature are given in the manuscript, and are also available upon reasonable request from the author.

## 5 Acknowledgements

This research has been supported by the European Research Council through the European Union’s Horizon 2020 research and innovation program under ERC Advanced Grant 788195 (FORMKIN). I thank Jade Mikalauskis for assistance with data compilation, and I acknowledge with gratitude the teaching of the late Dr. John L. Gill of Michigan State University.

## 6 Generative AI usage

Generative AI was not used in the preparation of this paper.

## Appendices

### A Some useful facts about conditional expectations and variances

Some things that are useful to know, or to recall if already known.

The conditional expectation of *ξ* conditional on the value of a random variable *R*

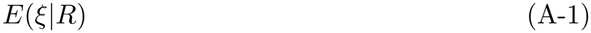

is itself a random variable. When a value of *R* is specified, e.g.,

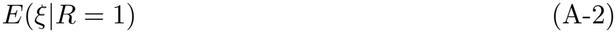

the conditional expectation stops being a random variable and becomes a number.

Because it is a random variable, one can take the expectation of the conditional mean over a mixing distribution of the values of *R*:

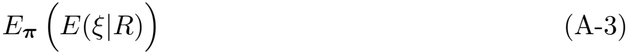

This conditional expectation is evaluated as an expectation over the possible values of *R*, with probabilities given by the distribution ***π***.

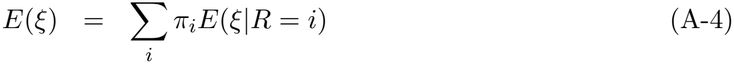

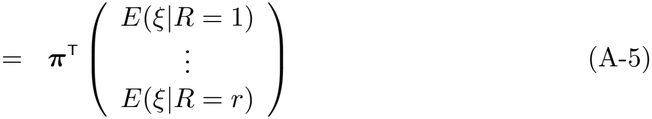

When *ξ* is conditional on two random variables, one of which is fixed, as in

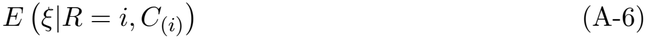

the result is a random variable with respect to *C*_(*i*)_, which becomes an actual quantity when a value is specified for *C*_(*i*)_. The matrix expressions for means in this paper (e.g., equation 28), apply to vectors of values, not to the random variables of which those vectors are realizations.

### B A protocol for hierarchical variance partitioning

We conclude with a step-by-step protocol for the partitioning of variance in a two-level hierarchical study design.

1. Choose a population and define a set of groups within this population. Let us call these Regions, although they need not be.
2. Within each region define a set of sub-groups. Let us call these sub-groups Countries.
3. Define a mixing distribution ***π*** giving the proportional representation of each region within the population. The only restriction is that ***π*** must be a probability vector.
4. Within each region *i*, define a mixing distribution ***σ***_*i*_ giving the proportional representation of the countries within the region *i*. The ***σ***_*i*_ must be probability vectors.
5. Choose a demographic outcome of interest; call this variable *ξ*.
6. Calculate the mean and variance of *ξ* for each country. This might be done with a life table or with a Markov chain or Markov chain with rewards.
7. Collect these means and variances into vectors within each region:

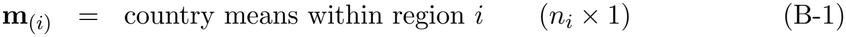

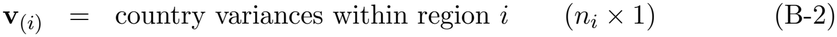

for *i* = 1, …, *k*
8. Calculate the mean of *ξ* within each region using equation (12). Collect these means into a vector **m**,

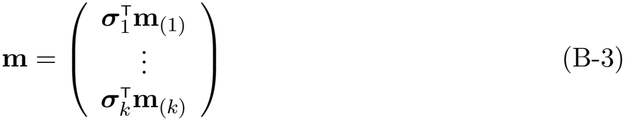
9. Calculate the overall population mean *E*(*ξ*) as a mean, over the mixing distribution ***π***, of the entries of **m**,

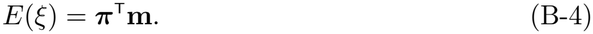
10. Calculate the variance of *ξ* within each region, including the within country and between-country components as in equations (19 and (20)
11. Collect the terms in these variances into two vectors,

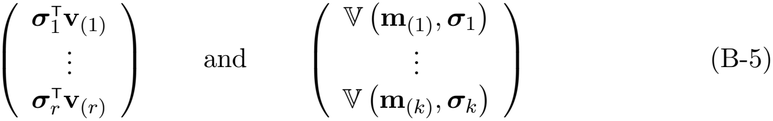

These appear in terms *C* and *B* in Equation (28).
12. Calculate the between-region variance as the variance, over the mixing distribution ***π***, of the entries of the vector of regional means, V (**m, *π***). This provides term *A* in Equation (28).
13. The result of this protocol is the global variance, the between-region variance, the between-country variance, and the within-country variance due to stochasticity.

### C Supplementary figures

This supplement contains the graphics for males, and for the flat mixing distribution, corresponding to Figures 5–7 in the main text.

#### C.1 Male, population-weighted mixing distribution

**Figure C-1:**
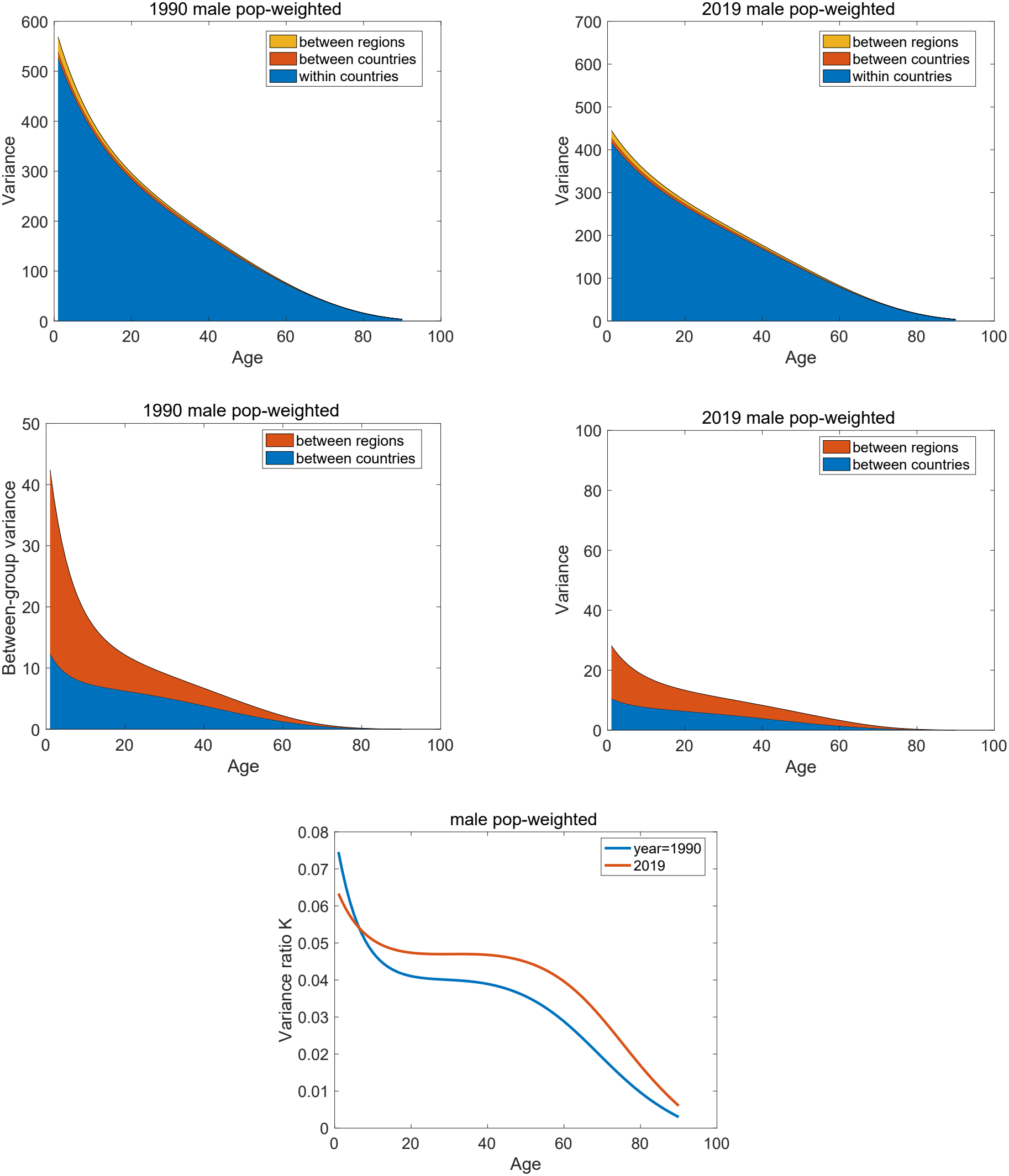
variance components and variance ratio, males, as a function of age in selected years

**Figure C-2:**
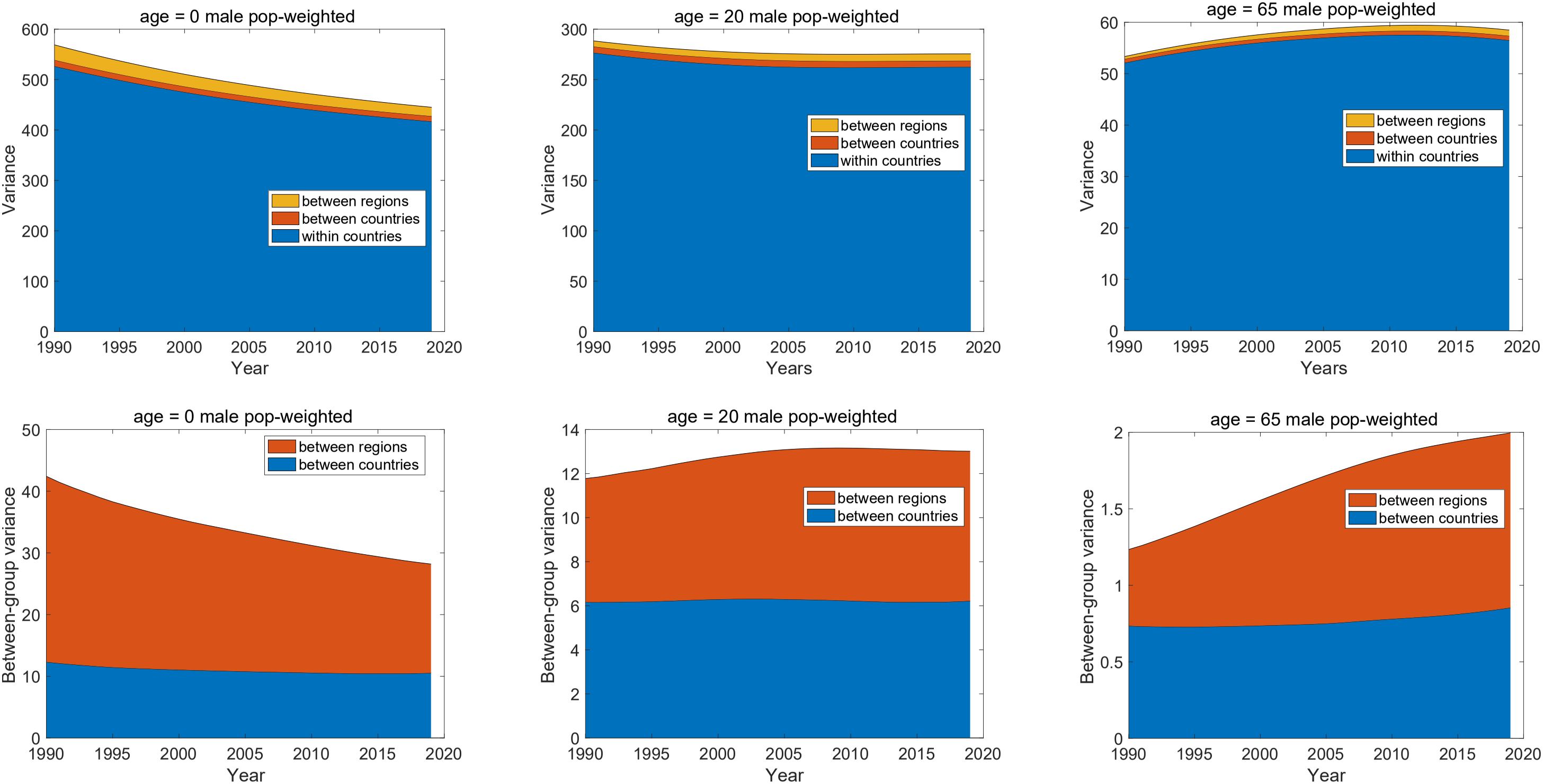
variance components for remaining male healthy longevity as a function of time, for selected ages (0, 20, 65).

**Figure C-3:**
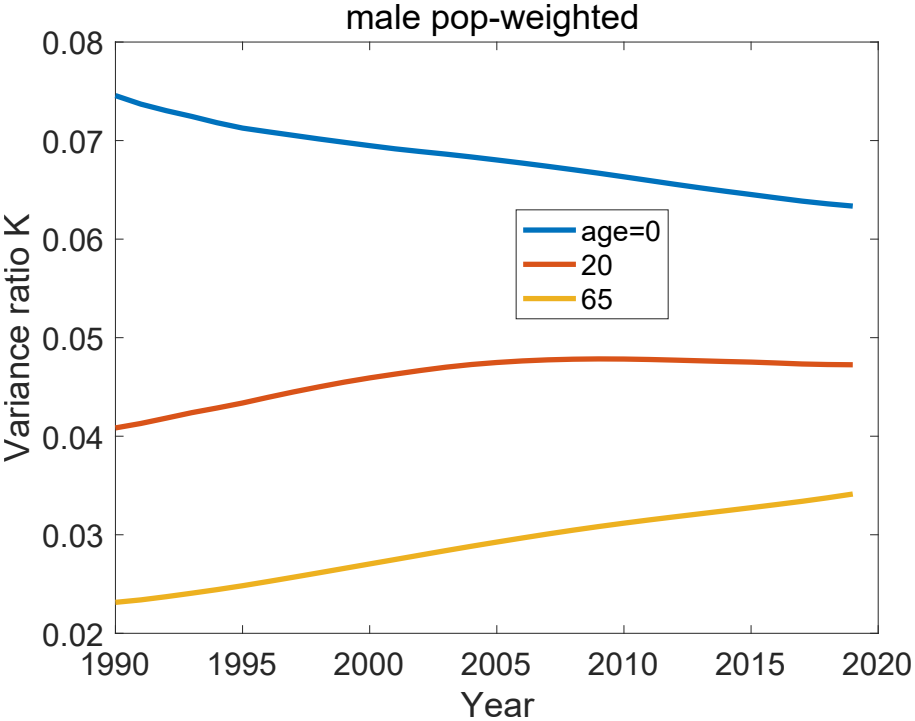
The variance *K* ratio for remaining male healthy longevity, as a function of time, for selected ages (0, 20, 65).

#### C.2 Female, flat mixing distribution

**Figure C-4:**
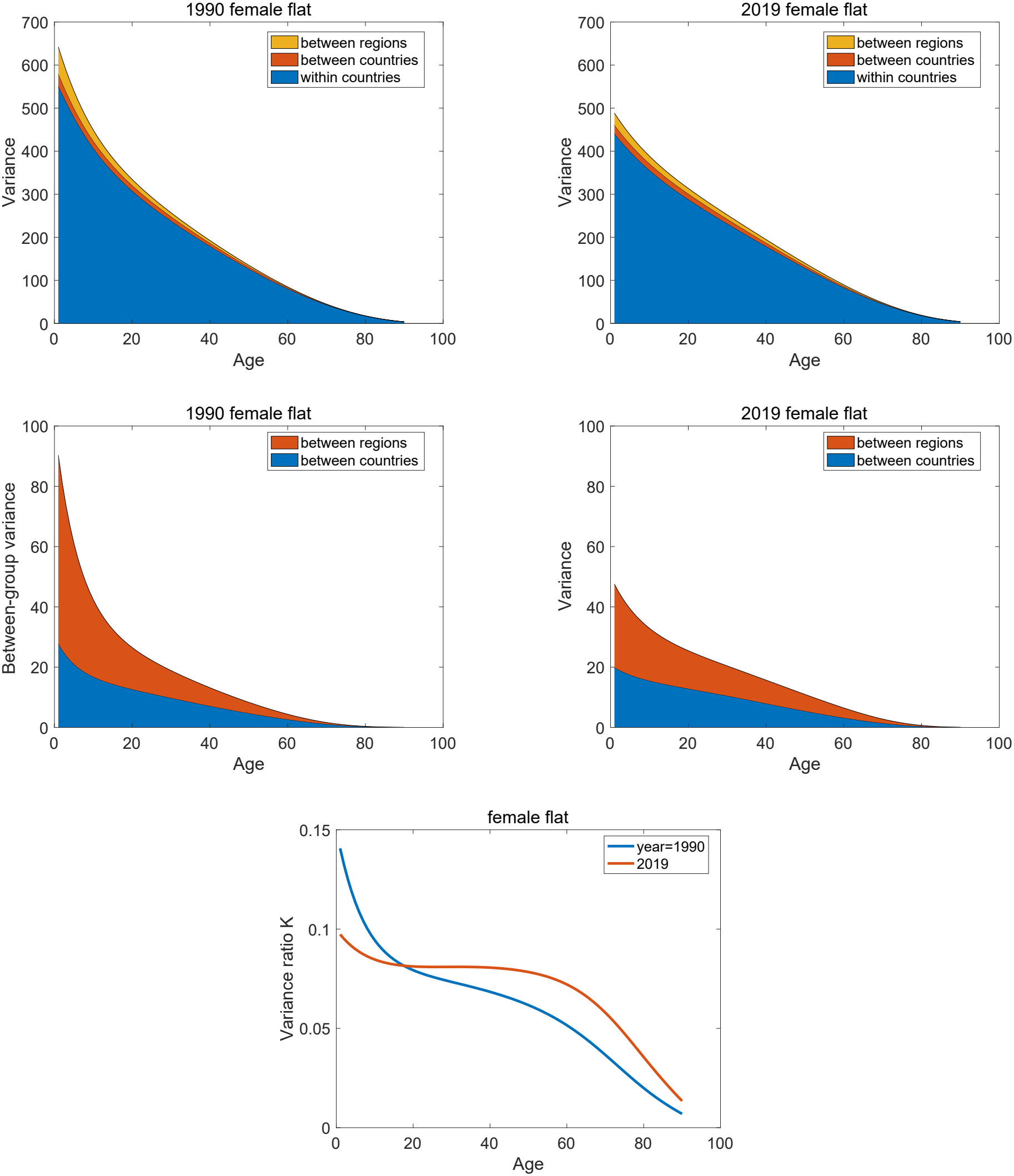
variance components and variance ratio, females, as a function of age in selected years

**Figure C-5:**
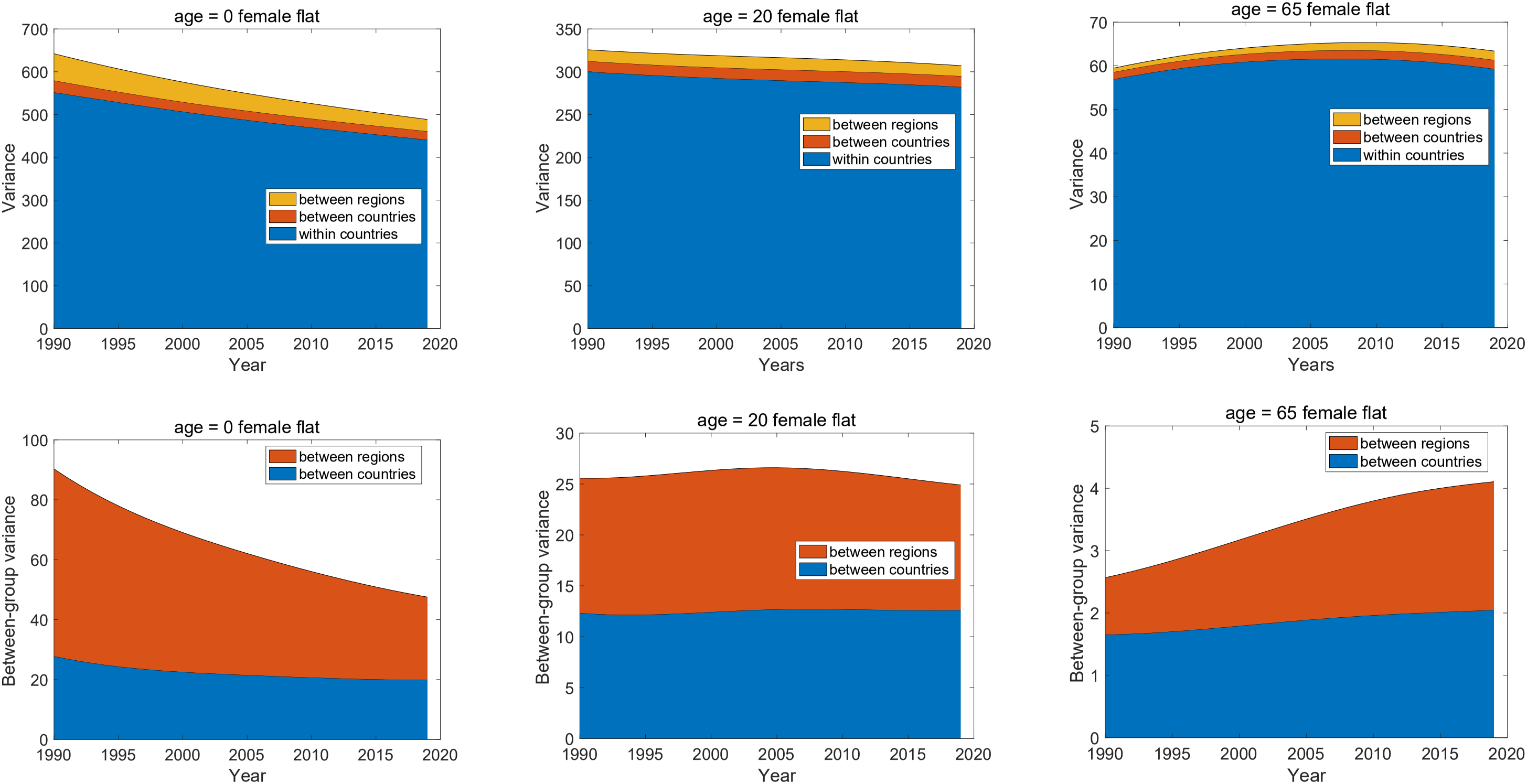
variance components for remaining female healthy longevity as a function of time, for selected ages (0, 20, 65).

**Figure C-6:**
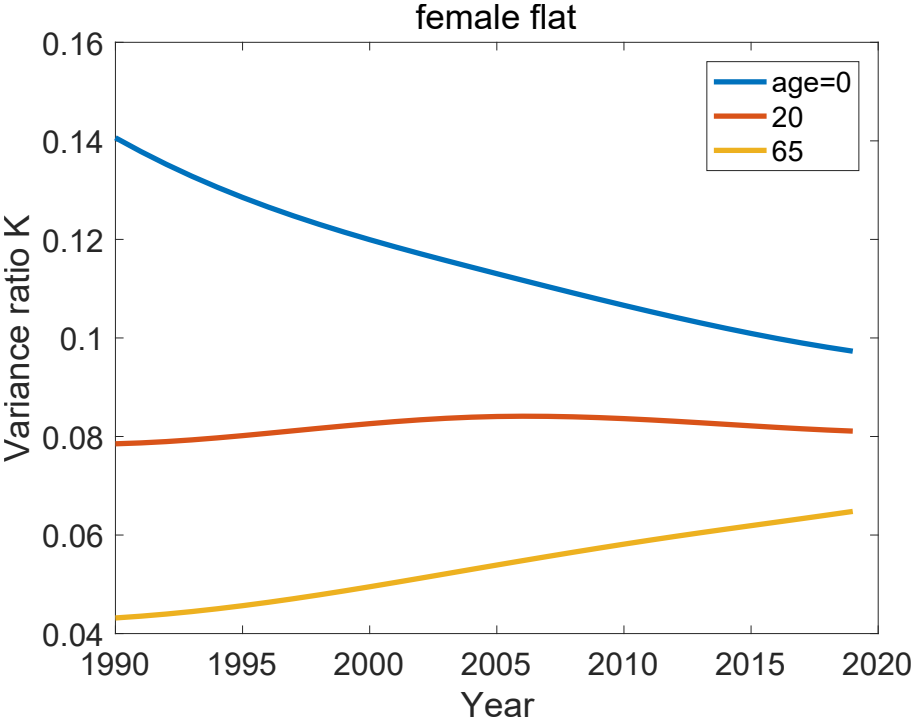
The variance ratio *K* for remaining female healthy longevity, as a function of time, for selected ages (0, 20, 65).

#### C.3 Male, flat mixing distribution

**Figure C-7:**
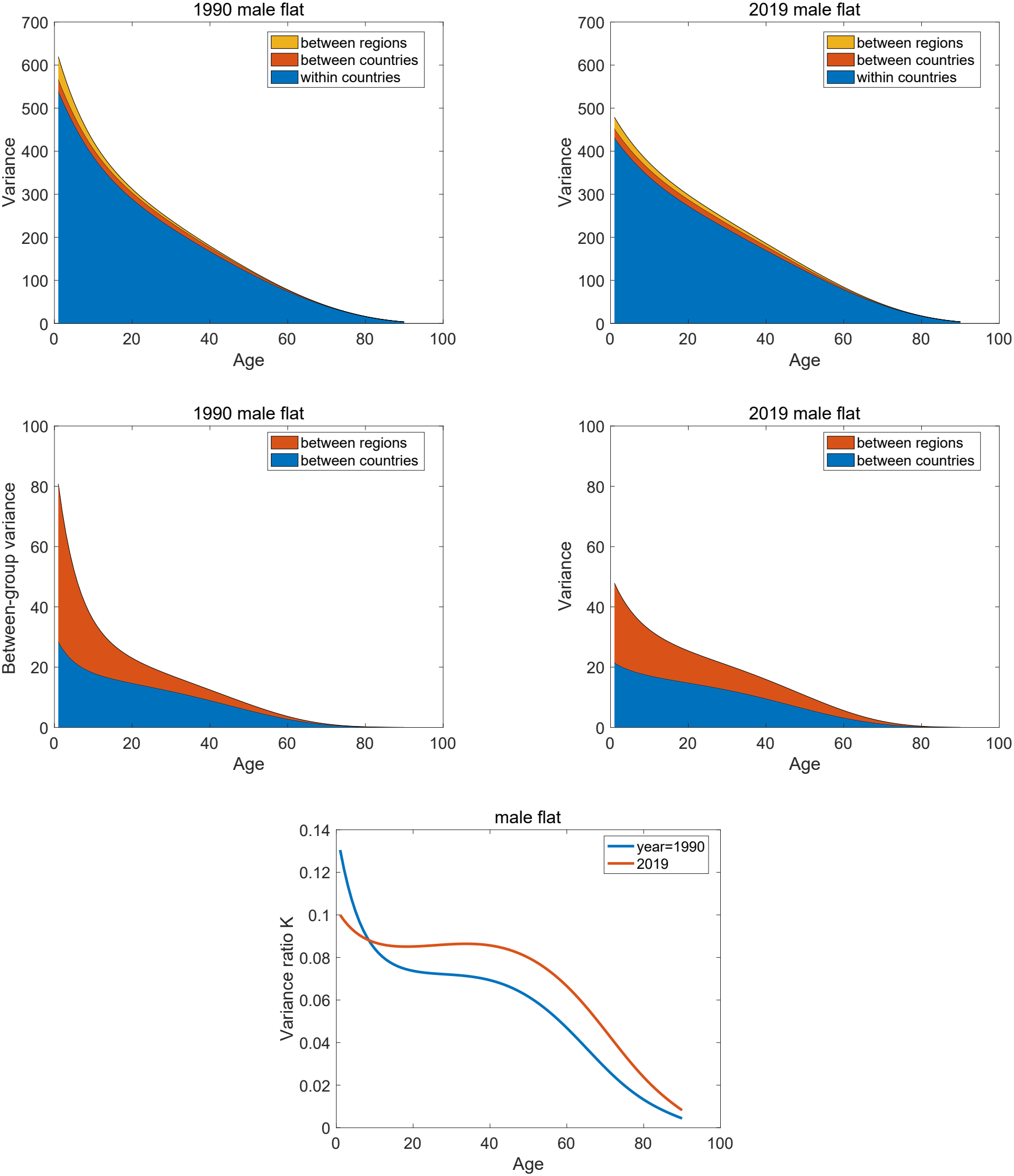
variance components and variance ratio, males, as a function of age in selected years

**Figure C-8:**
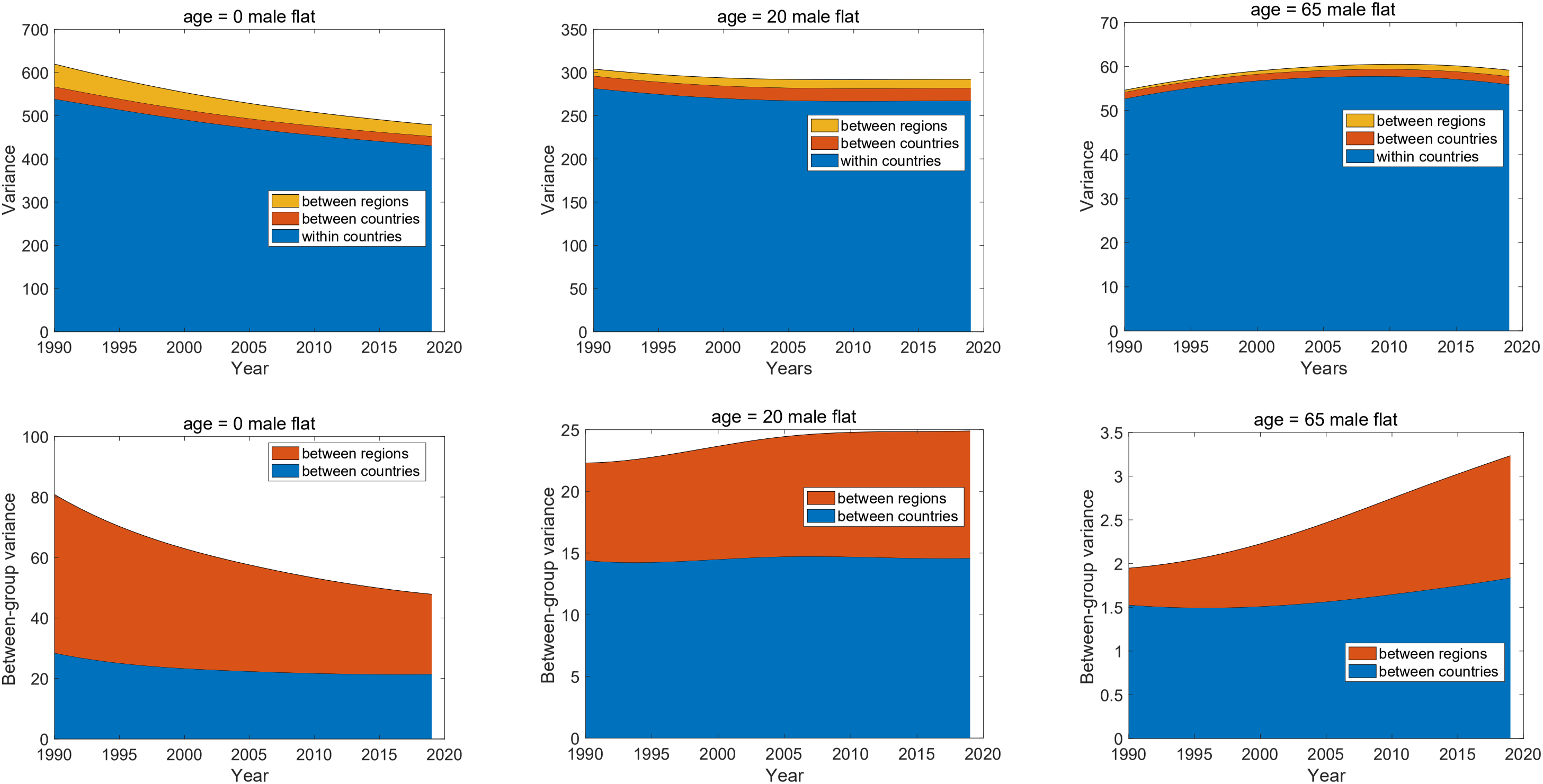
variance components for remaining male healthy longevity as a function of time, for selected ages (0, 20, 65).

**Figure C-9:**
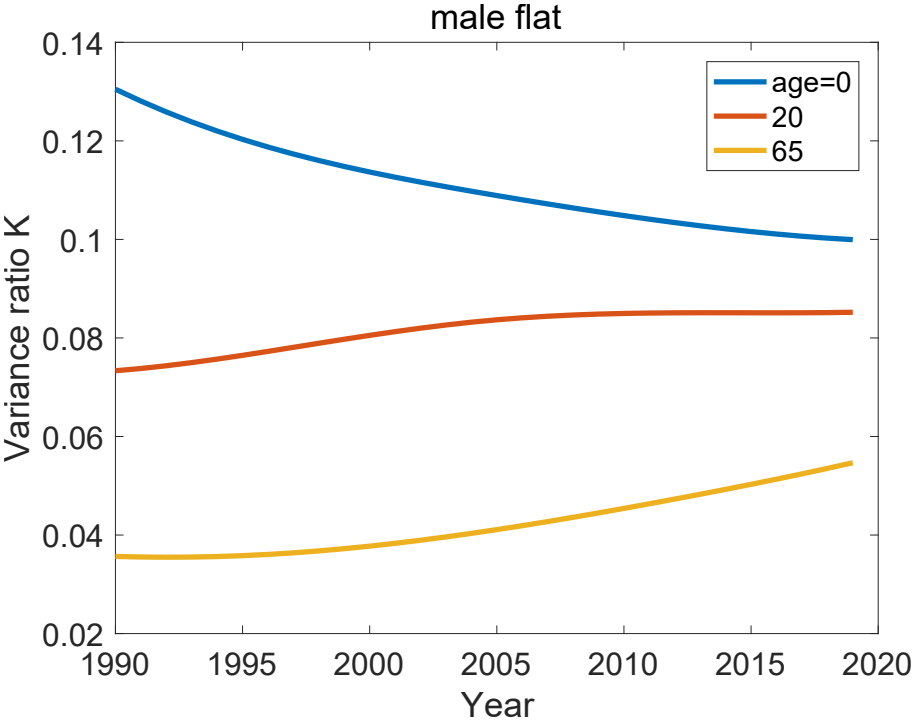
The variance ratio *K* for remaining male healthy longevity, as a function of time, for selected ages (0, 20, 65).

## Footnotes

1 I have been unable to find the line in Homer’s poem. It may have been created for the screenplay. If you know of a source, please let me know.

2 Even continuously distributed heterogeneity, like the gamma-distributed frailty in the gamma-Gompertz mortality model, can be discretized into groups (e.g., Hartemink, Missov, and Caswell, 2017) for numerical calculations.

3 Note that this argument applies to studies that use life tables, or Markov chains, to compute inequality in some demographic outcome. The (uncommon) situation when individual lifetimes are measured rather than calculated is fundamentally different; see Section 4.1.3.

4 Sub-Sarahan Africa, Northern Africa and Western Asia, Central and Southern Asia, Eastern and South-Eastern Asia, Latin America and the Carribean, Oceania, and Europe and Northern America; see https://unstats.un.org/sdgs/indicators/regional-groups/

5 https://blogs.worldbank.org/en/opendata/world-bank-country-classifications-by-income-level-for-2024-2025.

6 Corresponding graphs for males, and for the flat mixing distribution for both sexes, are given Supplementary Appendix C.

